# Neural mechanisms of disease pathology and cognition in young-onset Alzheimer’s Disease variants

**DOI:** 10.1101/2024.03.11.24304042

**Authors:** Seda Sacu, Catherine F. Slattery, Karl J. Friston, Ross W. Paterson, Alexander J.M. Foulkes, Keir Yong, Sebastian Crutch, Jonathan M. Schott, Adeel Razi

**Author notes:** **Correspondence to:** Adeel Razi Turner Institute for Brain and Mental Health, School of Psychological Sciences, Monash University, Australia Clayton VIC 3800.

## Abstract

Late-onset Alzheimer’s disease is consistently associated with alterations in the default-mode network (DMN)—a large-scale brain network associated with self-related processing and memory. However, the functional organization of DMN is far less clear in young-onset Alzheimer’s disease (YOAD). We assessed resting-state DMN effective connectivity in two common YOAD variants (i.e., amnestic variant (n = 26) and posterior cortical atrophy (n = 13) and healthy participants (n=24) to identify disease- and variant-specific connectivity differences using spectral dynamic causal modelling. Patients with the amnestic variant showed increased connectivity from prefrontal cortex to posterior DMN nodes relative to healthy controls, whereas patients with posterior cortical atrophy exhibited decreased posterior DMN connectivity. Right hippocampus connectivity differentiated the two patient groups. Furthermore, disease-related connectivity alterations were also predictive of group membership and cognitive performance. These findings suggest that resting-state DMN effective connectivity provides a new understanding of neural mechanisms underlying the disease pathology and cognition in YOAD.

## 1. Introduction

Alzheimer’s disease is a neurological condition characterized by progressive decline in cognitive functions. Young-onset Alzheimer’s Disease (YOAD, symptom-onset< 65 years) is a rare form of AD, which is associated with a more aggressive clinical course, greater tau burden, and prominent atrophy in posterior brain regions compared to more common late-onset AD^1–3^. Moreover, YOAD presents a heterogeneous clinical profile with variable cognitive and behavioral impairments^4^. Although most patients diagnosed with YOAD exhibit memory deficits—also called amnestic variant, a higher proportion of the patients present with atypical phenotypes characterized by other focal symptoms^2^. Posterior cortical atrophy (PCA) is the most common atypical phenotype dominated by visuoperceptual/visuospatial problems, parieto-occipital atrophy and relatively preserved episodic memory^5^. Despite the shared pathology, there are remarkable differences between the two YOAD phenotypes regarding grey matter volume, glucose hypometabolism and organization of large-scale brain networks^6,7^.

Growing evidence suggests that AD disrupts the communication between functional systems, which might further explain the decline in cognitive functions^8^. The default-mode network (DMN) is the most studied large-scale brain network in neurological conditions and associated with several cognitive processes such as autobiographical memory, future planning and self-awareness^9^. The activity of the DMN shows a striking correlation with the topography of amyloid β (Aβ)^10,11^ and tau^12,13^ deposition and is correlated with self-referential thoughts and episodic memory, which indicates that memory impairment identified in AD can be attributable to disrupted DMN connectivity ^10^. Taken together, these findings suggest that DMN connectivity is a promising and reliable brain phenotype to understand disease state as well as cognitive processes in AD.

Previous literature has consistently shown that patients with late-onset AD exhibited decreased local connectivity in the DMN ^14,15^ and decreased connectivity between the DMN regions, especially between posterior cingulate cortex and other default-mode network nodes ^16–19^. Compared to late-onset AD, a relatively small number of connectivity studies investigated differences in large-scale networks in patients with YOAD. These studies reported both increased^20^ and decreased^20–22^ functional connectivity between the DMN regions. Only a few studies investigated DMN connectivity in YOAD variants ^23,24^. Lehmann *et al.,* (2015) identified increased anterior DMN functional connectivity in amnestic AD and PCA compared to controls, whereas the other study found decreased DMN functional connectivity in the PCA variant compared to controls^24^. However, none of these studies examined directed (i.e., effective) connectivity in the DMN in patients with YOAD or YOAD variants.

In this work, we aimed to investigate effective connectivity between the DMN nodes in YOAD variants (i.e., amnestic AD and PCA) and healthy age-matched controls using dynamic causal modelling (DCM). DCM brings methodological advantages, which allows new insights into understanding disease pathophysiology and underlying neural mechanisms. Unlike functional connectivity methods, DCM can infer the direction of influences that nodes in network exert over each other (i.e. causal relationships in the context of the model) as well as the valence of the influence (i.e. inhibitory or excitatory signaling). Moreover, DCM can also infer self-connections, reflecting sensitivity of one region to inputs from other regions. Given the shared pathological mechanism between YOAD and late-onset AD, we think our methodology would also provide insights into disease state and cognition in late-onset AD. We hypothesized that patients with YOAD would show dominantly decreased connectivity in the DMN compared to healthy controls. In addition, based on structural and functional differences between the two AD variants^6,7^, we expected to see group differences in the DMN connectivity between the two patient groups, especially in posterior DMN regions and hippocampus. Furthermore, there is a scarce literature on behavioral correlates of connectivity alterations. To fill this gap, we investigated whether effective connectivity within DMN can predict cognitive performance and disease age-onset.

## 2. Methods

### 2.1. Participants

The case-control study included forty-five patients with probable AD^21^ with symptom onset < 65 years and 24 age-matched healthy individuals. The participants were recruited between 2013 and 2015 from Cognitive Disorders Clinic at the National Hospital for Neurology and Neurosurgery, London, United Kingdom. Patients were classified based on their leading symptoms as having typical amnestic ^25^ or atypical (PCA) AD phenotype^26^. Patients with an autosomal dominant mutation (n=1) or other atypical AD phenotypes (n = 3; two patients with logopenic progressive aphasia and one patient with behavioural/dysexecutive AD phenotype) were excluded from the study. Furthermore, two patients were excluded from further analyses due to excessive head motion (> 3 mm in translation or 3 degrees in rotation parameters) calculated based on six motion parameters obtained after realignment. The final sample included 26 patients with amnestic AD, 13 patients with PCA and 24 healthy controls (See Table 1 for sample characteristics). The study was approved by the National Hospital for Neurology and Neurosurgery Research Ethics Committee. All participants provided written informed consent.

### 2.2. Cognitive measurements

All participants underwent an extensive neuropsychology test battery measuring the severity of cognitive impairment (Mini-Mental State Examination [MMSE] ^27^), episodic memory (Recognition Memory Test [RMT] ^28^), numeracy and literacy respectively (Graded Difficulty Arithmetic [GDA] ^29^) and Graded Difficulty Spelling Test [GDST] ^30^], visuospatial and visuoperceptual performance (Visual Object and Spatial Perception [VOSP]^31^ battery) and speed of processing and executive functions (Delis-Kaplan Executive Function System [DKEFS]^32^). All cognitive measurements were continuous. For all scales, higher scores indicated higher cognitive performance.

### 2.3. Data acquisition and preprocessing

The data were acquired on a Siemens Magnetom Trio (Siemens, Erlangen, Germany) 3T MRI scanner using a 32-channel phased array head coil. Structural images were obtained using a sagittal magnetization-prepared rapid gradient echo (MP-RAGE) three-dimensional T1-weighted sequence (TE = 2.9 ms, TR = 2200 ms, TI = 900 ms, flip angle = 10°, FoV = 282 × 282 mm, 208 slices with 1.1×1.1×1.1 mm voxels). Functional images were obtained using an asymmetric gradient echo echo-planar sequence sensitive to blood oxygen level-dependent contrast (36 contiguous interleaved slices, TE= 30 ms, TR = 2200 ms, flip angle = 80°, FoV = 212 × 212 mm, voxel size = 3.3 × 3.3 × 3.3 mm). In total, 140 volumes were acquired during the resting state scan.

Preprocessing steps were performed using SPM12 (version 7771). The initial five volumes were discarded to allow for equilibration of the magnetic field. Slice acquisition dependent time shifts were corrected per volume. All volumes were realigned to the first volume using a six-parameter (rigid body) linear transformation to correct within subject head motion. The resulting images were then spatially normalised to a standard brain template in the MNI coordinate space. Data were resampled to 3-mm isotropic voxels and spatially smoothed using a 6-mm full-width half-maximum (FWHM) Gaussian kernel. Six motion parameters and time series from white matter and cerebrospinal fluid (i.e., first principal component of time series within the subject-level anatomical image obtained after segmentation) were used as nuisance regressors for physiological noise correction.

### 2.4. The regions of interest selection

Constrained independent component analysis (ICA) was performed to identify the network of interest using the Group ICA of fMRI Toolbox (https://trendscenter.org/software/gift/). Constrained ICA is a semi-blind dimension reduction method that identifies an independent component in subject data, which matches with the supplied spatial references ^33^. This approach features two main advantages. First, it eludes problems with subjective component selection ^31^. Second, the resulting components have higher signal-to-noise ratio than the standard ICA approaches.

A pre-existing template for DMN (dorsal DMN)^34^ was used as a spatial reference for constrained ICA. A one-sample t-test was performed to obtain group-level peak coordinates for regions of interest (ROIs). Based on the t-test results (p < 0.05, family-wise error (FWE) corrected), we selected following ROIs for subsequent DCM analysis: medial prefrontal cortex (mPFC; x=-6, y=53, z=17), posterior cingulate cortex (PCC; x=6, y=-55, z=29), bilateral angular gyrus (ANG; left: x=-48, y=-67, z=35; right: x=48, y=-61, z=29) and bilateral hippocampus (HPC; left: x=-24, y=-25, z=-13; right: x=27, y=-25, z=-10). These coordinates are very similar to the anatomical locations reported by a coordinate-based meta-analysis of DMN in healthy adults and AD patients ^35^. ROI time series were defined as the first principal component from all voxels within in a 8 mm radius sphere centered on the group maximum of each node ^36^. Subject-specific whole brain mask and MNI-based ROI masks were used to ensure that regional responses were based on voxels within the anatomical boundaries of the brain and specific ROI.

### 2.5. Data analysis

#### 2.5.1. Statistical analysis

Statistical analysis was performed in SPSS 24 (SPSS Inc., Chicago, Illinois, USA). A nominal significance threshold was set at *p*=0.05. Chi-squared tests were performed to identify if sex and handedness were equally distributed across groups. A one-way ANOVA was performed to compare age, education, neuropsychological test scores, and mean head motion between groups. Non-parametric ANOVA test (Kruskal-Wallis) was used for the same purpose when parametric assumptions were not met. Two-sample t tests were then performed to compare two patient groups in terms of disease duration and age at onset.

#### 2.5.2. Independent component analysis

After identifying the subject-specific DMN component using constrained ICA, we conducted a one-way ANOVA test in SPM to see if there are group differences in DMN component (i.e., spontaneous fluctuations in BOLD signal) between amnestic AD, PCA and healthy controls. We here focused only the ROIs that we used in the DCM model and reported cluster-level FWE corrected results.

#### 2.5.3. Spectral dynamic causal modelling

##### 2.5.3.1. Subject Level

The spectral DCM implemented in the SPM12 (DCM 12.5, revision 7497) was performed to estimate effective connectivity during resting-state (see Supplementary Material S2 for methodological details and priors on parameters). The spectral DCM estimates resting-state connectivity by fitting observed complex fMRI cross spectra. It finds the best effective connectivity among the hidden neuronal states that explains functional connectivity among haemodynamic responses ^36–38^. Here, we set a fully connected model (6 x 6 = 36 connection parameters) to compare all possible nested models within the network^39^. After the model estimation, we performed diagnostics to check the quality of the DCM model fitting to ensure that model inversion was successful^40^. All participants had more than 60% explained variance by the model (M = 89.5, SD = 3.9).

##### 2.5.3.2. Group Level

A parametric empirical Bayes (PEB) analysis was then performed at the group level to estimate the group mean and group differences in effective connectivity^39^. PEB in this setting entails a general linear model (GLM) of between-subject (random) effects that best explain the within-subject (DCM) estimates of connectivity. Bayesian Model Reduction (BMR) was used to prune connection parameters from the full PEB model until there was no further improvement in model-evidence (based on log-evidence or free energy). The parameters of reduced models were then averaged and weighted by their model evidence by Bayesian Model Averaging (BMA) to account for any uncertainty over reduced models: i.e., the parameters under each reduced model were averaged after weighting by the evidence (a.k.a., marginal likelihood) of the model under which they were estimated.

Due to the group differences in head motion, we included mean framewise displacement in the analysis as a covariate of no interest at the between the subject level. All inferences were made using a posterior probability criterion of 95% (equivalent of a strong evidence) for each connection. In other words, the model evidence or marginal likelihood of a model with a particular parameter was 20 times greater than the corresponding model in which the parameter was removed. Since PEB is a multivariate Bayesian GLM, in which all the connectivity parameters are fitted at once to optimize the model evidence, no correction for multiple comparisons is required in contrast to a frequentist approach. See Figure 1 for the analysis pipeline.

**Figure 1.**
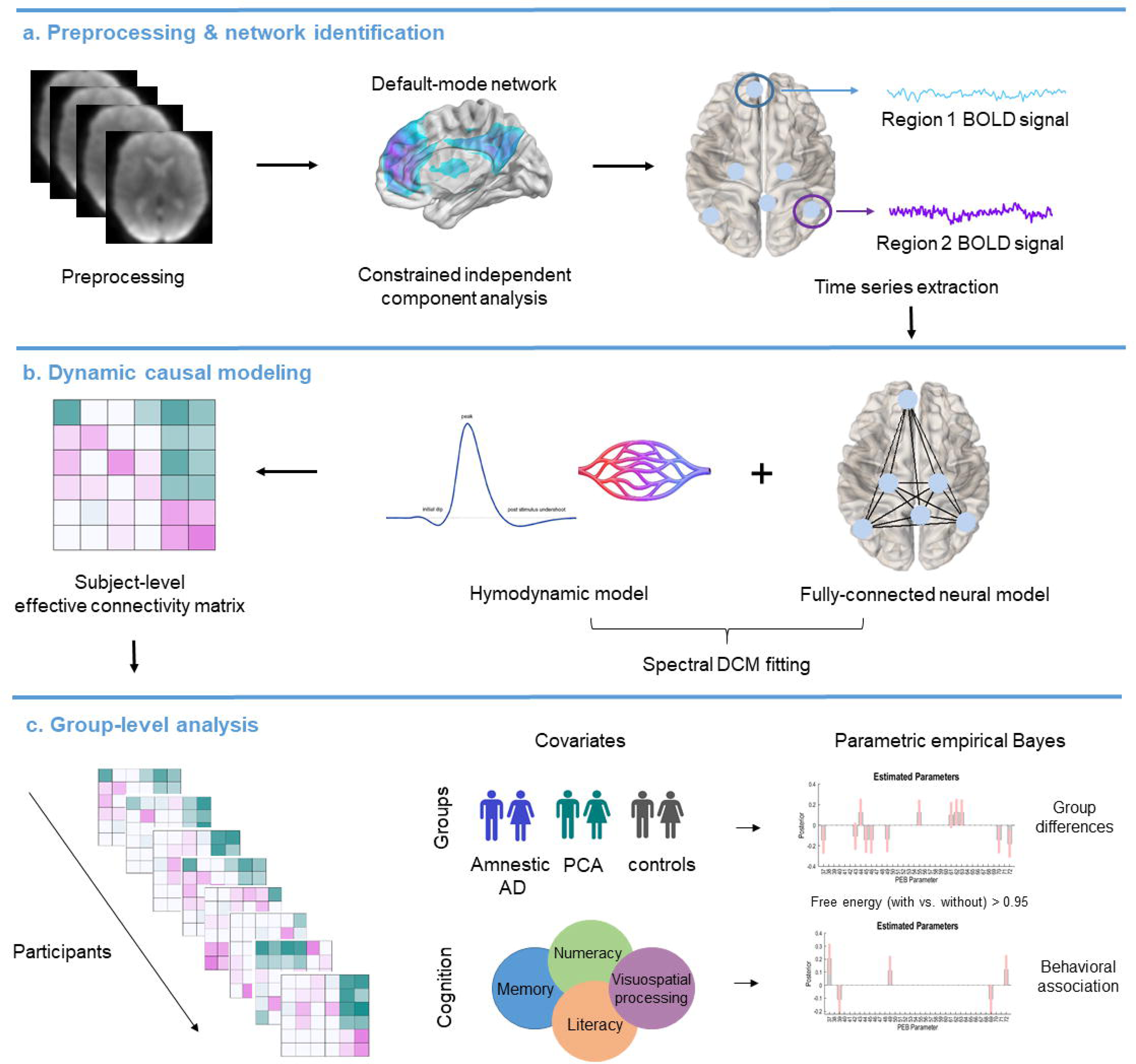
Graphical summary of the analysis pipeline.

#### 2.5.4. Cognitive Performance

We investigated the associations between effective connectivity parameters and cognitive performance using the PEB framework. To present a composite score reflecting cognitive performance, we first conducted a principal component analysis using MMSE, RMT, GDA, GDST, VOSP, and DKEFS sum scores (See Supplementary Material S3 for details). Only one component—which explained 68% of the variance—was identified and used for the analysis. All inferences were made using a posterior probability criterion of 95% (equivalent of a strong evidence) for each connection.

#### 2.5.5. Leave-one-out cross validation

We used leave-one-out cross-validation approach implemented in the PEB framework to test whether an effective connectivity parameter can predict group membership (e.g., amnestic AD, healthy controls) and cognitive performance. We first examined if the connectivity parameters, which we found to be significant to characterize group differences, can predict group membership. We then tested whether cognitive performance can be predicted by effective connectivity parameters across participants (n=63), independently from group differences.

#### 2.5.6. Sensitivity analysis

Here, we used a DMN component called dorsal DMN as a spatial reference—or constraint—for the ICA analysis, since it covers most reliable DMN regions, such as PCC, medial PFC and angular gyrus ^35^ as well as hippocampus—an important brain region for AD pathology ^41^. Although DMN is mostly treated as a homogenous network, several studies have identified distinct components within DMN ^34,42,43^. Furthermore, some studies have shown that certain DMN components were differentially altered in AD ^15,23,44^. Therefore, we also included another DMN component called ventral DMN^34^ to see if our results are specific to the chosen component or YOAD pathology. The dorsal DMN component contained midline structures such as PCC and mPFC and located more anteriorly—akin to anterior DMN, whereas ventral DMN included larger involvement of posterior midline structures such as PCC/dorsal precuneus and relatively less and lateralized involvement of frontal regions (Supplementary Fig. S1&S2). The same analysis pipeline as dorsal DMN was applied for the sensitivity analysis (see Supplementary Material S5).

## 3. Results

### 3.1. Sample Characteristics

Groups were matched in terms of age, sex, education, and handedness. However, there was a significant group difference in head motion quantified by using frame-wise displacement ^45^. Patients with amnestic AD had significantly higher head motion than healthy controls (*t(48)* = -0.32, p = 0.03). Patient groups did not differ in terms of disease severity and duration.

As expected, patient groups had significantly lower scores in all neuropsychological tests than the control group and composite cognition score obtained from principal component analysis (p < 0.001, See Table 1). Patients with PCA had lower scores in VOSP than the patients with amnestic AD (p < 0.05). Patient groups did not significantly differ in other neuropsychological tests and in composite cognitive score. There was no significant group difference in MMSE, disease onset, and disease duration between the two patient groups.

Earlier disease age-onset was related to lower cognitive performance indexed by the composite cognition score (r=0.43, p=0.006). No such association was found between disease duration and cognitive performance.

### 3.1. Independent component analysis

A component representing dorsal DMN was identified across participants using constrained ICA (Figure 2). Within this component, we found a group difference between amnestic AD and healthy controls. YOAD patients with amnestic phenotype showed lower PCC activation compared to healthy controls (*p* < 0.05, cluster-level FWE-corrected; Supplementary Fig. S3). However, this result did not survive at whole-brain corrected level. No group difference was found for any ROI within the ventral DMN component.

**Figure 2.**
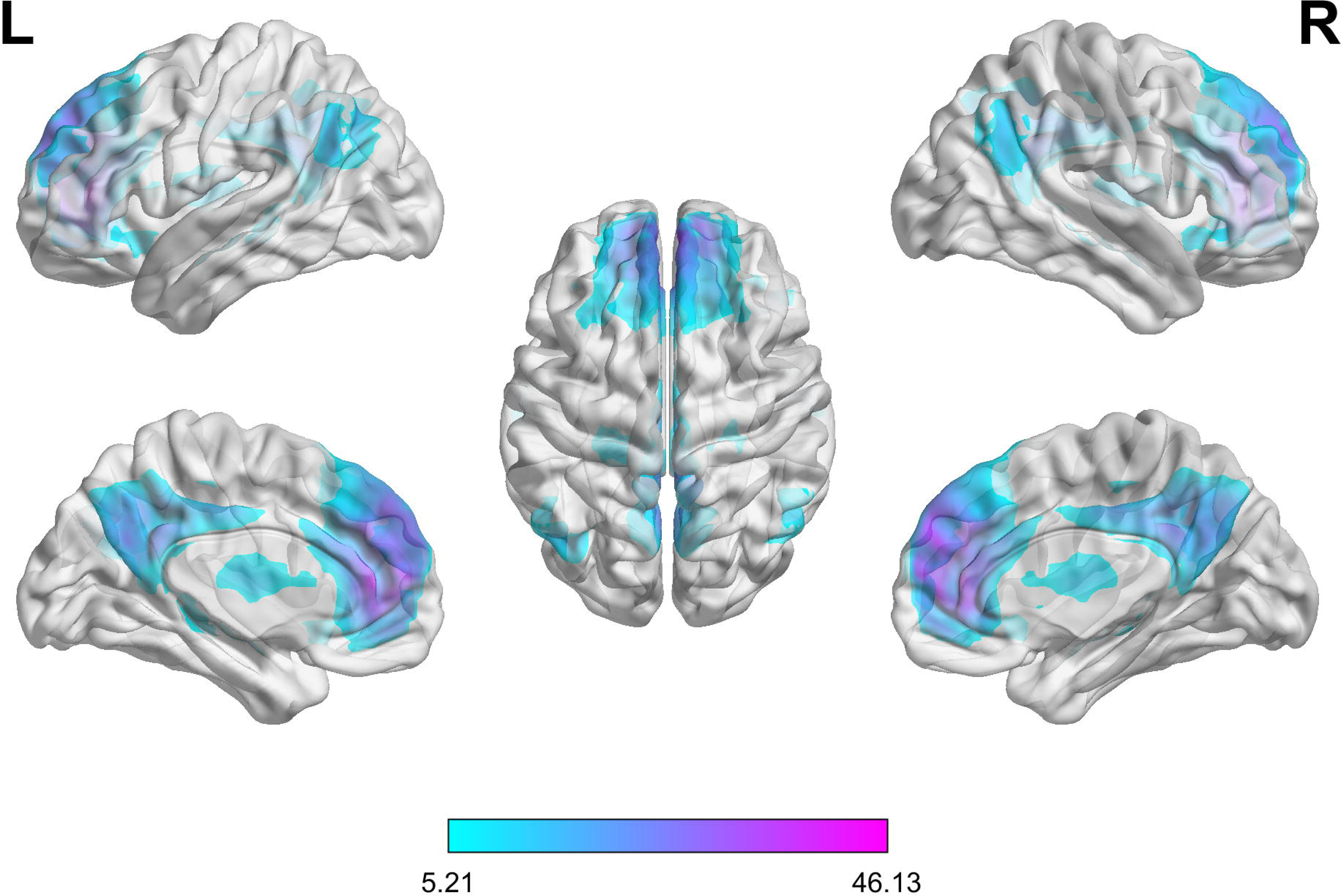
Default-mode network component identified using constrained independent component analysis (p <0.05, FWE-corrected at whole-brain).

### 3.2. Dynamic causal modelling

#### 3.2.1. Group means

Group specific effective connectivity matrices for dorsal DMN are shown in Figure 3 (see Supplementary Fig. S5 for the ventral DMN component). Two types of connection between regions can be seen in Figure 3: excitatory and inhibitory (extrinsic or between-region) connections. An excitatory connection means that the activity of the source region leads to increases in the activity of the target region, whereas an inhibitory connection means that the activity of the source region leads to decreases in the activity of the target region ^46^. In addition, a self-connection (e.g., PCC ➔ PCC) refers to the recurrent activity in brain region and is always inhibitory. The self-connections regulate a region’s response to its inputs; i.e., they provide gain control in the model, akin to inhibitory inter-neurons – and consequent excitation-inhibition balance – in the brain. A positive self-inhibitory connection indicates a more inhibited region, which is less responsive to the input, while a negative self-inhibitory connection indicates a less inhibited region resulting in disinhibition to afferent input^40^.

**Figure 3.**
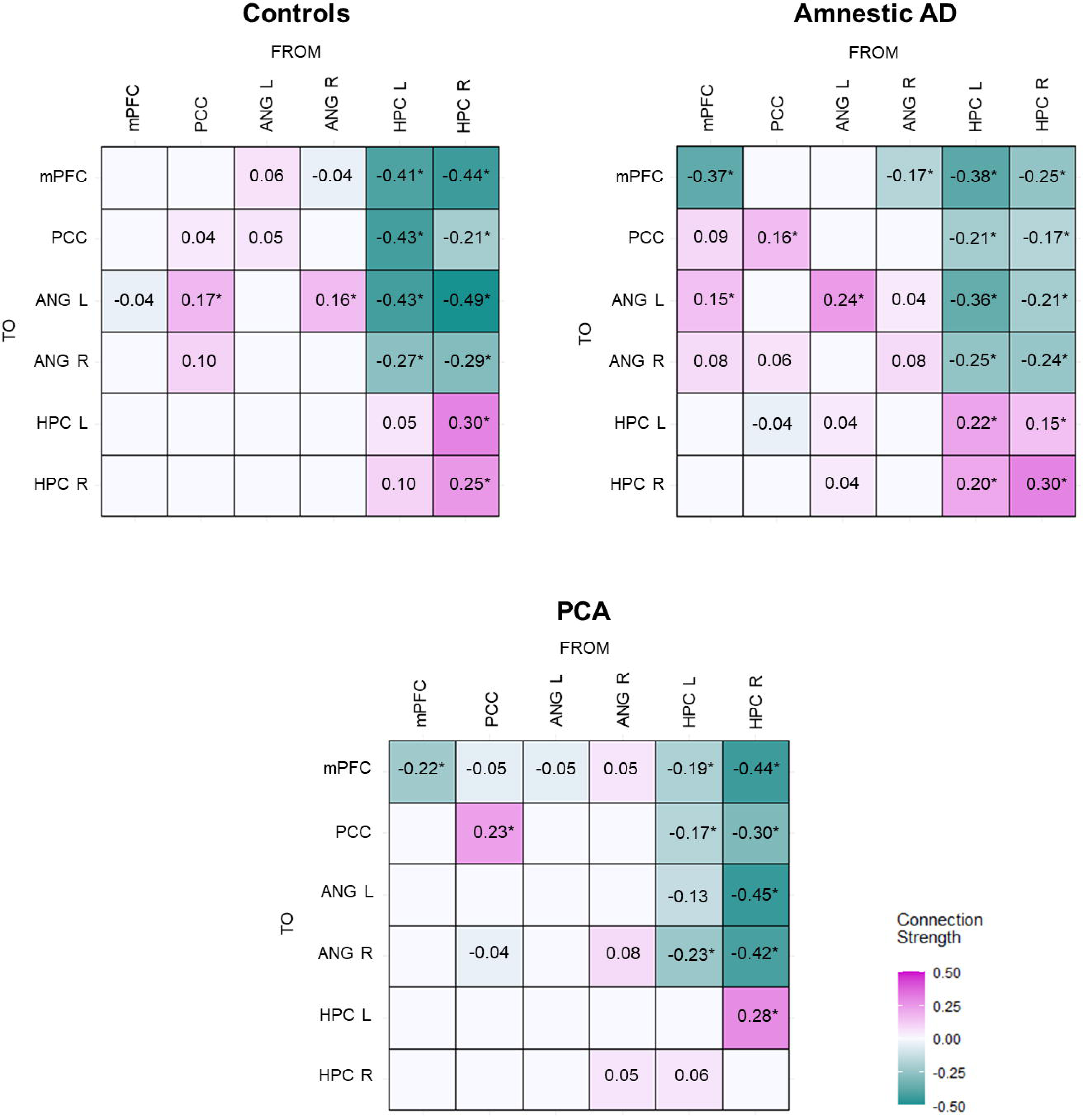
Mean effective connectivity within the DMN. Magenta color represents excitatory connections, whereas green color represents inhibitory connections. Self-connections should be interpreted as inhibitory, irrespective of the color. Connections with strong evidence [i.e., *posterior probability* > 0.95] are marked with an asterisk. Abbreviations: ANG, angular gyrus; HPC, hippocampus; mPFC, medial prefrontal cortex; PCC, posterior cingulate cortex.

The groups showed a mixed pattern of excitatory and inhibitory connections within DMN. However, outgoing connections from HPC to other DMN regions were inhibitory across all groups, while interhemispheric HPC connections were excitatory, which is in line with a recent study investigating effective connectivity in healthy elderly and demented patients^47^. Missing entries correspond to redundant connections that have been removed following Bayesian model reduction (that did not differ from their prior expectation).

#### 3.2.2. Group differences

##### 3.2.2.1. Amnestic AD versus healthy controls

Compared to the healthy controls, patients with amnestic AD showed lower mPFC self-connection indicating higher input sensitivity of the region in the disease group (Figure 4). In addition, patients with amnestic AD had higher connectivity from mPFC to posterior DMN regions (i.e., PCC and bilateral ANG) than the healthy controls. The patient group exhibited excitatory connections from mPFC to posterior DMN regions, which were missing in the healthy group. Contrary to the mPFC self-connection, left ANG self-connection was higher in the patient group indicating the lower input sensitivity of the region. Additionally, patients with amnestic AD variant showed lower inhibitory connectivity from left HPC to PCC and from right HPC to mPFC and left ANG than the healthy controls.

**Figure 4.**
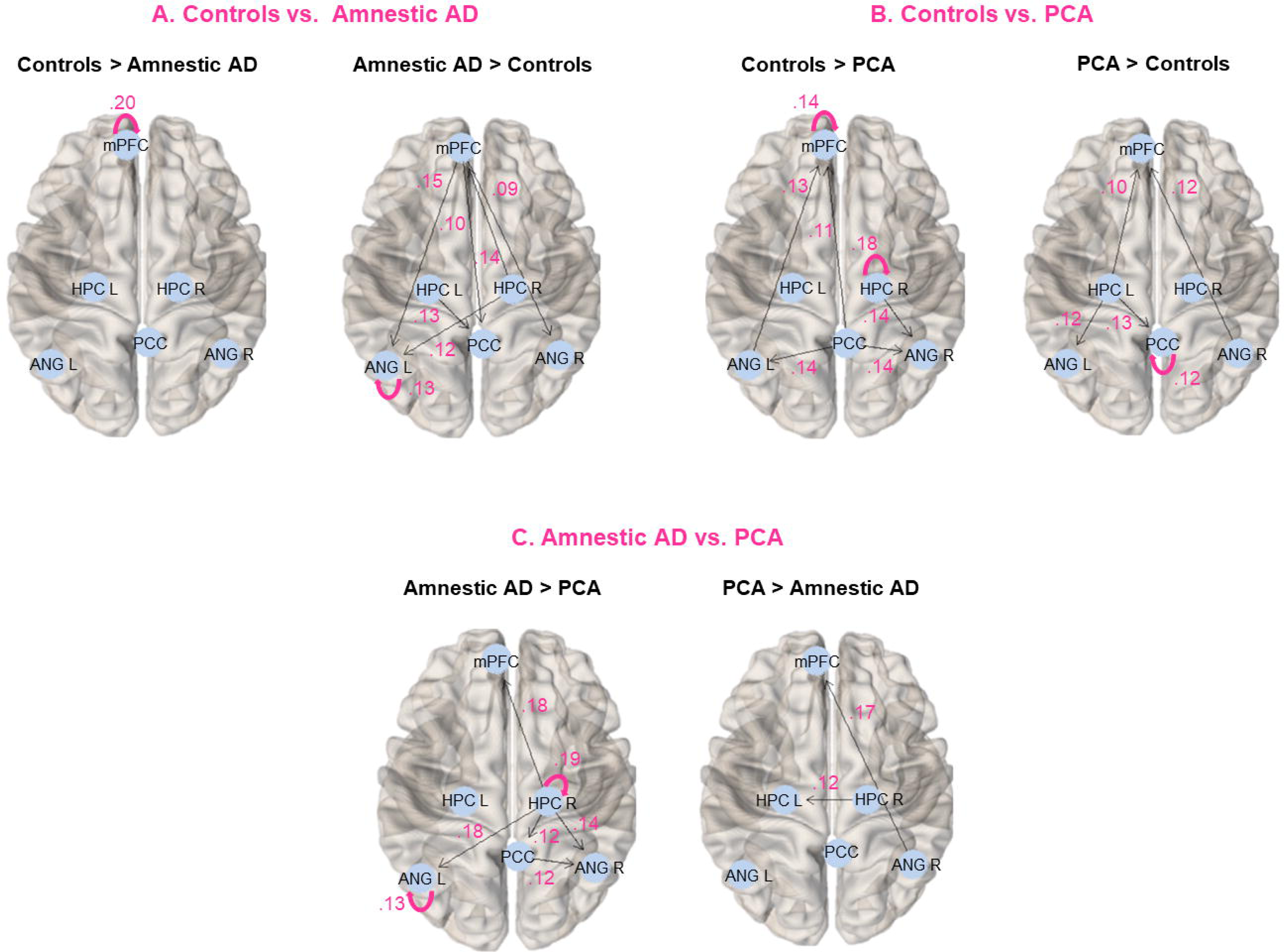
Group differences in DMN effective connectivity. Results were mapped on the brain using brainconn (https://github.com/sidchop/brainconn) implemented in R. Abbreviations: AD, Alzheimer’s disease; ANG, angular gyrus; HC, healthy controls; HPC, hippocampus; mPFC, medial prefrontal cortex; PCA, posterior cortical atrophy; PCC, posterior cingulate cortex.

Similarly, we identified higher input sensitivity of right middle frontal gyrus, higher connectivity from right middle frontal gyrus to other DMN regions (left middle frontal gyrus and bilateral angular gyrus) and lower input sensitivity of right ANG in patients with amnestic AD compared to healthy controls in the ventral DMN (Supplementary Fig. S6).

##### 3.2.2.2. PCA versus healthy controls

Similar to the patients with amnestic AD variant, patients with PCA exhibited lower mPFC self-connection (i.e., higher input sensitivity) compared to the healthy controls (Figure 4). The PCC-self connection was higher (i.e., lower input sensitivity) in the patients with PCA than the healthy controls. In addition, patients showed lower connectivity from PCC to other DMN regions (e.g., mPFC and bilateral ANG). Indeed, connections from PCC to bilateral ANG were excitatory in the healthy group, whereas they were missing or weakly inhibitory in the patients with PCA. Additionally, the patient group showed lower inhibitory connectivity from left HPC to mPFC, PCC and left ANG, lower right HPC self-connection, and higher inhibitory connectivity from right HPC to right ANG compared to the healthy controls.

Similarly, we identified higher input sensitivity of right middle frontal gyrus together with increased connectivity from right middle frontal gyrus to bilateral ANG and higher input sensitivity of right parahippocampal gyrus in patients with PCA compared to the healthy controls in the ventral DMN (Supplementary Fig. S6).

##### 3.2.2.3. Amnestic AD versus PCA

Patients with amnestic AD had lower inhibitory right HPC connectivity to other DMN regions, and lower excitatory connectivity from right HPC to left HPC than the patients with PCA variant (Figure 4). Moreover, right HPC self-connection was higher in the amnestic AD group indicating lower sensitivity of the right HPC to input from other regions. Additionally, patients with amnestic AD exhibited lower left ANG self-connection, higher connectivity from PCC to right ANG (i.e., excitatory in the amnestic group) and lower connectivity from right ANG to mPFC (i.e., inhibitory in the amnestic group) compared the PCA group.

Similarly, we found that patients with amnestic AD exhibited lower input sensitivity of right parahippocampal gyrus and lower inhibitory connectivity from right parahippocampal gyrus to other DMN regions compared to the patients with PCA in the ventral DMN (Supplementary Fig. S6).

### 3.3. Cognitive Performance

Higher cognitive performance was related to higher mPFC and right HPC self-connections (i.e., more inhibited, less sensitive to input), lower connectivity from mPFC and right HPC to left ANG and higher connectivity from left ANG to mPFC (Figure 5).

**Figure 5.**
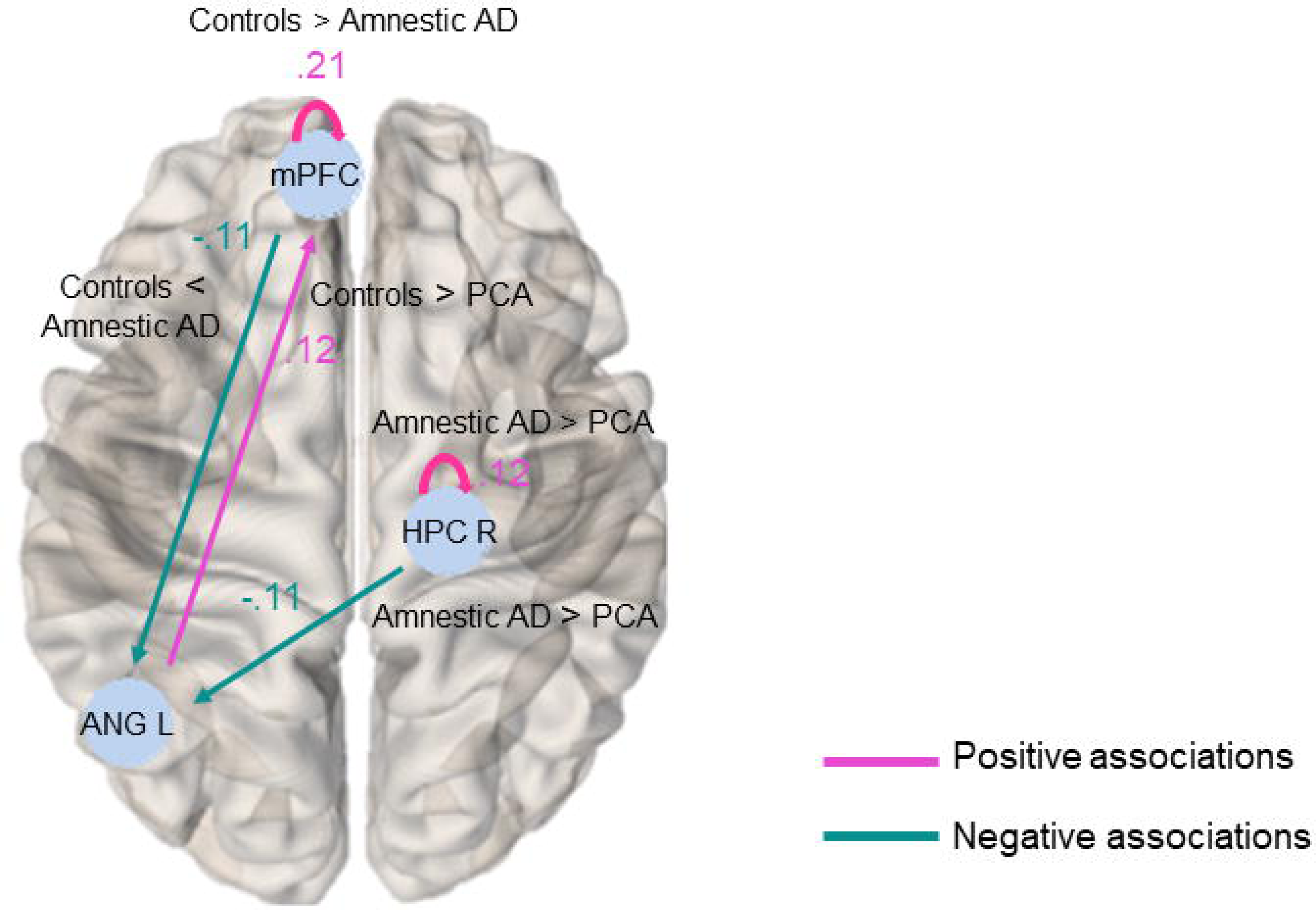
Associations between cognitive performance and effective connectivity. Magenta color represents positive associations (i.e., higher connectivity strength-higher cognitive performance), whereas green color represents negative associations (i.e., higher connectivity strength-lower cognitive performance). Group differences related to the cognitive performance-effective connectivity associations were also reported in the figure. Abbreviations: ANG, angular gyrus; HPC, hippocampus; mPFC, medial prefrontal cortex.

### 3.4. Leave-one-out cross validation

Table 2 shows effect of each connectivity parameters in predicting group membership and cognitive performance.

#### 3.4.1. Group membership

Effective connection from mPFC to PCC distinguished amnestic AD from controls (r=0.25, p < 0.037). Five connectivity parameters (ANG L ➔ mPFC, ANG R ➔ mPFC, ANG R ➔ HPC R, PCC ➔ ANG R, HPC R ➔ HPC R) predicted patients with PCA (PCA vs. controls) (r=[0.31-0.36], all p < 0.05). Similarly, five connectivity parameters (mPFC ➔ PCC, PCC ➔ ANG R, HPC R ➔ ANG R, PCC ➔ HPC L, HPC R ➔ HPC R) were large enough to distinguish YOAD variants (amnestic vs. PCA) (r=[0.27-0.4], all p < 0.05).

#### 3.4.2. Cognitive Performance

Four connectivity parameters (mPFC ➔ mPFC, ANG L ➔ mPFC, ANG L➔ PCC, and HPC R ➔ ANG L) were predictive for cognitive performance across participants (r=[0.22-0.25], all p < 0.05).

## 4. Discussion

We here investigated effective connectivity within the DMN in two YOAD variants and healthy elderly to identify disease and variant-specific network alterations in the DMN. Our results indicated that both patient groups exhibited increased mPFC sensitivity to afferent inputs and a reduced inhibitory influence from left HPC to PCC, in comparison to healthy participants. Patients with amnestic AD uniquely showed excitatory connectivity from mPFC to posterior DMN nodes compared to healthy participants, whereas patients with PCA exhibited decreased connectivity from PCC to other DMN nodes and reduced inhibitory influence from left HPC to other DMN nodes. The connectivity of right hippocampus differentiated the patient groups. Importantly, the disease- and variant-specific connectivity alterations were able to distinguish groups and predict cognitive performance, suggesting that DMN effective connectivity provides important insight into understanding disease pathology and cognition.

### 4.1. Hyperexcitability in amnestic AD variant

Reduced DMN functional connectivity is one of the most replicated findings in late-onset AD ^14,48–50^. In line with this, we hypothesized that the patients with amnestic AD would show decreased DMN connectivity compared to healthy participants. On the contrary, we found that patients with amnestic variant had unique excitatory efferent connections from mPFC to posterior DMN nodes, which were missing in healthy controls and lower inhibitory influence from hippocampus to other DMN nodes. The studies conducted with YOAD patients reported both decreased ^20,22,51^ and increased ^20,23,52^ functional connectivity in patients. In addition, there is no one-to-one mapping between functional and effective connectivity. Indeed, they can be profound quantitative and qualitative differences between the two methods ^38^. This follows from the fact that a change in one effective (directed) connection can change the correlations throughout the network—and therefore cause distributed changes in functional connectivity (See the group mean functional connectivity in Supplementary Fig. S4 for comparison with the effective connectivity estimates in Figure 2).

Moreover, increased functional connectivity in DMN can be component specific ^15,23^. To exclude the possibility that our results were driven by the component choice, we repeated the same analysis using the ventral DMN. Independent from the component, we here identified increased input sensitivity of frontal regions and increased inter-regional effective connectivity from frontal regions to posterior nodes. Additionally, this pattern was not seen only in the amnestic AD variant but also in the PCA. Taken together, these results suggest that increased frontal cortex connectivity is related to disease pathology rather than the heterogeneity of DMN.

However, the meaning of increased connectivity is not clear. Some authors interpret increased connectivity as an early compensatory mechanism in response to the loss of functional capacity^53,54^. However, damage to a neural system may also result in increased connectivity^55^. This phenomenon has been documented in several neurodegenerative disorders including Parkinson’s disease, multiple sclerosis, and mild cognitive impairment ^54–57^. Although hyperconnectivity can be adaptive in the short-term, chronic hyperconnectivity may make the neural system vulnerable to secondary pathological processes ^55^.

Importantly, brain regions showing Aß deposition are located within DMN^10^. Thus, Aβ-induced hyperexcitability can be a plausible explanation for the disinhibition findings in the current study. Animal model of AD suggests that elevated Aß deposition can induce neuronal hyperexcitability and trigger epileptiform activity in the hippocampal and cortical network^58,59^. Indeed, epileptic seizures are prevalent in AD, especially with younger onset^60,61^. Although Aß is widely used to explain neural hyperexcitability, the effect of Aß on functional connectivity is not clear. Some studies showed that Aß leads to decreases in functional connectivity, while others showed the other way around ^62^.

Tau protein might also play a role in the increased DMN connectivity in YOAD^63^. Increased anterior-posterior connectivity was found to be associated with increases in frontal tau deposition over time ^64,65^. However, some other studies reported decreased connectivity related to tau deposition^62^. Unfortunately, we did not collect any neuropathological data to examine how protein deposition can be related to network alterations in YOAD. Therefore, this explanation remains speculative. However, DCM can be a promising method to combine with neuropathological data, in future, to investigate whether enhanced excitatory or reduced inhibitory signalling is related to protein deposition in AD. Especially increased effective connectivity from the mPFC to posterior nodes might reflect AB-dependent dysfunctional signaling in early stages of dementia, which can also help us to understand disease progress in late-onset AD.

### 4.2. Mixed connectivity pattern in PCA

We hypothesized that the patients with PCA would exhibit reduced effective connectivity between DMN nodes compared to healthy participants. Our hypothesis was partially confirmed for the PCA variant. Healthy participants exhibited excitatory connections from PCC to mPFC and bilateral ANG indicating network integrity of DMN. However, patients with PCA either exhibited inhibitory influences from PCC to the other nodes or lack of outgoing efferent PCC connections. The PCC was also less sensitive to inputs of other DMN regions in patients with PCA than the healthy controls. These results were in line with previous studies reporting decreased DMN functional connectivity in patients with PCA ^24,66^. Moreover, several previous studies reported structural^5,66^, functional ^67^ and connectomic ^23,68^ parietal cortex abnormalities in patients with PCA.

However, it is important to note that patients with PCA did not show only decreased effective connectivity. The effective connectivity from right ANG gyrus to mPFC was weakly excitatory in patients with PCA, whereas the same connection was inhibitory in healthy elderly and patients with amnestic AD. In addition, we identified a reduced inhibitory influence from left HPC to mPFC, left ANG and PCC than the healthy controls, which can be interpreted as a disinhibitory effect. The latter also shows that network alterations were not restricted to parietal regions. Although medial temporal regions are relatively spared in PCA^5^, patients with PCA showed structural^7^ and connectomic^68^ abnormalities in the temporal cortex compared to healthy elderly. Taken together, although the connectivity of parietal regions plays a more important role in PCA, the network alterations are not limited to parietal regions.

### 4.3. Differential hippocampal connectivity in YOAD variants

Patients with amnestic AD had reduced inhibitory connectivity from right hippocampus to other DMN regions compared to the patients with PCA. Additionally, the right HPC was less sensitive to inputs of other DMN regions and had a reduced excitatory outgoing connection to the left HPC in the amnestic AD variant. Our replication analysis revealed similar results for the right parahippocampal gyrus in the ventral DMN. These results confirm that brain regions related to episodic memory functions are affected more in amnestic AD than PCA.

HPC is an important brain region for memory and learning and one of the earliest affected brain regions in AD ^41^. Although both patient groups show grey matter abnormalities in the HPC, the differences are less in patients with PCA compared to patients with amnestic AD^7,69^. Therefore, group differences in hippocampal connectivity between the two YOAD variants can be expected. However, it is not clear why we found the group differences, specifically in the right hemisphere. A previous study reported higher total right hippocampus volume in patients with PCA than the patients with amnestic AD^67^. However, another study found left-sided group differences in hippocampus subfield volumes between the two AD variants^7^.

### 4.4. Predicting group membership and cognitive performance

Here, we showed that effective connectivity parameters related to group differences were helpful in predicting group membership. These results align with a recent study showing that resting-state DMN effective connectivity can predict dementia incidences before disease-onset in a relatively larger sample^47^, which supports the clinical usefulness of DMN effective connectivity in neurodegenerative disorders.

Moreover, YOAD provides a unique opportunity to understand cognition. We here found that earlier disease-onset was associated with poorer cognitive performance independent from disease duration. The effect of various connection parameters (e.g., mPFC self-connection, ANG L ➔ mPFC and HPC R to ANG L) was large enough to predict both disease status and cognitive performance. The overlapping mechanism between the disease effect and cognition indicated intertwined nature of these processes. For this reason, studying these processes together can help us to better understand the complex relationship between the brain and behavior.

### 4.5. Limitations

This study has several limitations that should be noted. First, our sample included only amnestic and PCA variants of AD. Investigating disease and variant-specific network alterations in other atypical AD variants such as logopenic progressive aphasia and behavioral AD could furnish new insights. Additionally, although comparable to the previous studies, our sample included fewer PCA cases (n=13), which can lead to reduced power in statistical analyses. Second, we did not measure Aβ or tau levels across the brain. We suggest that future studies should include neuropathological measurements to explain pathology behind the changes in the functional brain architectures. Third, the current study was cross-sectional. Longitudinal studies are needed to show how effective connectivity within and between networks change with disease progression. Fourth, network alterations in YOAD is not limited to DMN and distinct network abnormalities can be linked to specific YOAD phenotypes^70^. Therefore, future studies should investigate effective connectivity in other large-scale networks as well. Fifth, our cross-validation analysis should be considered as preliminary due to small sample size and should be interpreted carefully.

### 4.6. Conclusion

We here investigated disease- and variant-specific alterations in the DMN connectivity in two common YOAD variants using spectral DCM. Our results indicated that the resting-state DMN effective connectivity is a sensitive measure to detect disease- and variant-specific alterations in YOAD. However, since it was the first effective connectivity study in patients with YOAD variants, further research is needed to replicate these results.

## Acknowledgements

The authors thank all study participants for their contribution in scientific advance.

## Author Contributions

JMS and SC conceptualized the study. JMS obtained the funding. CFS, RWP, AJMF, and KY collected the data. AR supervised the study. SS conducted data analysis, visualized the results and wrote the first draft of the manuscript. All authors critically reviewed and edited the final version of the manuscript.

## Data Availability

The data that support the findings of this study are available from the corresponding author, upon reasonable request.

## Code Availability

The code to reproduce the findings is available from the corresponding author, upon reasonable request.

